# Cortical physiological arousal following termination of absence seizures

**DOI:** 10.64898/2026.10.20.26366013

**Authors:** Ayushe Sharma, Aya Khalaf, Sándor Beniczky, Tracy A. Glauser, Hal Blumenfeld

## Abstract

Although the brain state preceding and during absence seizures has been well-characterized, the postictal brain state remains largely unexplored due to the prevailing assumption that absence seizures have minimal or no postictal changes. To address this gap, scalp EEG recordings from 55 patients with 3-4 Hz spike-wave discharges across four multi-site cohorts were analyzed. Spectral power was measured relative to each seizure’s preictal baseline, and the postictal period was compared to a time-matched preictal window. Rather than an immediate return to baseline after seizure offset, a significant broadband increase in EEG power was observed in the gamma (>25 Hz) and delta (<4 Hz) frequency bands, followed by a gradual decline over the next 15 s to 30 s. In contrast, no comparable change preceded the seizures. This suggests that recovery from an absence seizure is an active process with a characteristic electrophysiological signature consistent with cortical activation and arousal. Thus, postictal arousal may facilitate exit from the seizure state, and with further work could serve as an electrophysiological marker and therapeutic target for recovery in absence epilepsy.

## Introduction

Brief, generalized 3 to 4 Hz spike-wave discharges (SWDs) are a defining feature of childhood and juvenile absence epilepsy. Clinically, absence seizures are subtle and often terminate as abruptly as they begin. Following several seconds of behavioral arrest, individuals typically resume activity without noticeable interruption. Interestingly, absence seizures occur most commonly in quiet, waking states of decreased arousal and are less frequent during times of increased arousal and attentional vigilance.^1; 2^ This may be related to mechanisms of absence seizure generation, which depend on abnormal, synchronous thalamocortical rhythms, promoted by states of decreased arousal.^3^ In addition, prominent sensory stimuli can sometimes interrupt absence seizures, which has been proposed to involve a transient increase in arousal.^4; 5^ We therefore hypothesized that spontaneous termination of absence seizure episodes may also involve an increase in arousal, reducing the susceptibility to abnormal thalamocortical rhythms as seizures end.

To investigate whether there is evidence for increased cortical physiological arousal with absence seizure termination, we examined electroencephalography (EEG) data at the end of SWDs in patients with absence epilepsy. We analyzed power across frequency bands, with broadband increases in gamma (>25 Hz) and delta (<4 Hz) power serving as markers of overall cortical activation, as demonstrated in prior work.^6-8^ Although physiological changes prior to absence seizures have been studied previously,^2; 9-12^ the immediate post-discharge period remains relatively unexamined. Investigation of the postictal EEG revealed an unexpected, brief, broadband power increase, most prominent in the initial seconds following seizure termination and at both higher (>25 Hz) and lower (<4 Hz) frequencies. These findings support the concept of an active, arousal-like emergence rather than a passive return to baseline at the time of absence seizure end.

## Materials and Methods

### Dataset descriptions and participants

Scalp EEG data from four studies, all previously analyzed by Springer *et al*. (2022), were included.^13^ Two studies were conducted at Yale: 1) a generalized epilepsy study of patients ≥15 years, recorded with a 128-electrode cap at 1000 Hz,^14^ and 2) a childhood absence epilepsy study of patients aged 6 to 19 years, recorded with a high-density 256-lead cap at 500 Hz.^9^ The third study used cup electrodes at 500 or 256 Hz, positioned according to the 10-20 system or the updated International Federation of Clinical Neurophysiology array to collect data at the Danish Epilepsy Center.^15^ The fourth was a multi-center study on childhood absence epilepsy organized by Cincinnati Children’s Hospital Medical Center, using data from 31 locations with a standard 10-20 montage collected at 256 Hz.^16^ Included participants exhibited bilateral 3 to 4 Hz SWDs on an otherwise normal EEG background, indicating idiopathic generalized epilepsy. Participants with any seizure types aside from typical absence seizures were excluded.

### EEG preprocessing and analysis

The onset and offset times of each SWD were marked by expert review. A modified version of Springer *et al*.’s processing pipeline^13^ was implemented in EEGLAB (Swartz Center for Computational Neuroscience, UCSD)^17^ and MATLAB R2023b (MathWorks). This involved visual inspection to remove bad channels (flat, noisy, or artifact-laden), resampling to a common frequency of 500 Hz, applying a 1 Hz high-pass finite-impulse-response filter to remove drift, removing line noise with CleanLine,^18^ and then applying a common-average reference. For each seizure, spectral power was analyzed over a preictal window (45 to 1 s before onset) and a postictal window (1 to 45 s after offset), excluding the ictal discharge. We required successive seizures to be ≥ 16 s apart in the preictal period and ≥ 11 s apart in the postictal period to include those periods in the analysis. We also excluded all preictal (up to 10s) postictal (up to 10s) and ictal data from another seizure occurring in the preictal or postical 45s windows. Preictal and postictal portions of each seizure were segmented into 1 s epochs, and any non-seizure epochs with voltage magnitude > 1.75 × mean voltage magnitude of the first 2 s of the corresponding SWD were discarded to remove high-amplitude movement and muscle artifact.

EEG signal power analysis before and after SWDs was based on an approach used previously.^10^ EEGs were converted to spectrograms using a single-taper, Hann-windowed short-time Fourier transform (5 s window, 0.5 s step, 1 to 100 Hz) with MATLAB’s spectrogram function, and power was averaged across all available channels. Preictal data were analyzed from -45 to -1 s before the SWD onset and postictal data were analyzed from 1 to 45 s after the SWD offset, with the ictal interval excluded. Analyses of long-term state changes in power before and after seizures were conducted by calculating mean power at each frequency from 45 to 1 s baseline before the initiation of each seizure, then normalizing power values in all 1 s time epochs by dividing by the mean baseline power for each corresponding frequency. The resulting values were log-transformed to express power in dB and smoothed with a 10-second moving average. Per-seizure power values in dB were then averaged across all seizures. Power calculations for statistical analysis in the frequency bands of interest (delta 1 to 3 Hz, theta 4 to 7 Hz, alpha 8 to 13 Hz, beta 14 to 24 Hz, and gamma 25 to 100 Hz) were calculated in the same way, except power values for each seizure were averaged over the relevant frequency ranges after baseline correction and log-transformation.

### Statistics

Data are presented as mean and standard error of the mean (SEM) across seizures. For each frequency band, power in dB calculated as already described was averaged over the postictal window (1 to 45 seconds) and compared to the matched preictal window (-45 to -1 seconds) for each seizure using a paired Wilcoxon signed-rank test (paired differences were non-normal; Lilliefors test), with paired Cohen’s d as the effect size. The five band p-values were corrected with the Benjamini-Hochberg false-discovery rate, and a band was significant at q < 0.05 with |d| ≥ 0.10.

## Results

### Study sample

The initial data sample was derived from cohorts described in detail for a previous study.^13^ Excluding seizures with insufficient preictal or postictal data (see Methods) led to a final sample of 232 seizures in 55 patients who had both preictal and postictal EEG data for analysis. Across the 55 patients, mean age was 10.1 ± 9.6 years (range 4.7 to 76 years; 37 female; 18 male) and mean seizure duration was 7.8 ± 6.5 seconds.

### Broadband postictal gamma and delta power increases

Before seizures, spectral power at all frequencies remained relatively unchanged over time (Figure 1A). However, in the postictal period, there was an increase in power in the first seconds after seizure offset, most prominently in the frequency ranges above 25 Hz (gamma), and below 4 Hz (delta) (Figure 1B). These increases gradually returned to baseline within approximately 15 to 30 seconds after seizure end (Figure 1B). Statistical analysis of power in the postictal (1 to 45 seconds) versus the matched preictal window (-45 to -1 seconds), showed significant postictal power increases in the delta band (+1.15 ± 0.27 dB, mean ± SEM; q = 0.019, Cohen’s d = 0.26) and in the gamma band (+1.34 ± 0.23 dB; q = 0.001, d = 0.33) (Figure 1C). Theta, alpha, and beta power did not differ significantly from the preictal baseline (Figure 1C).

**Figure 1.**
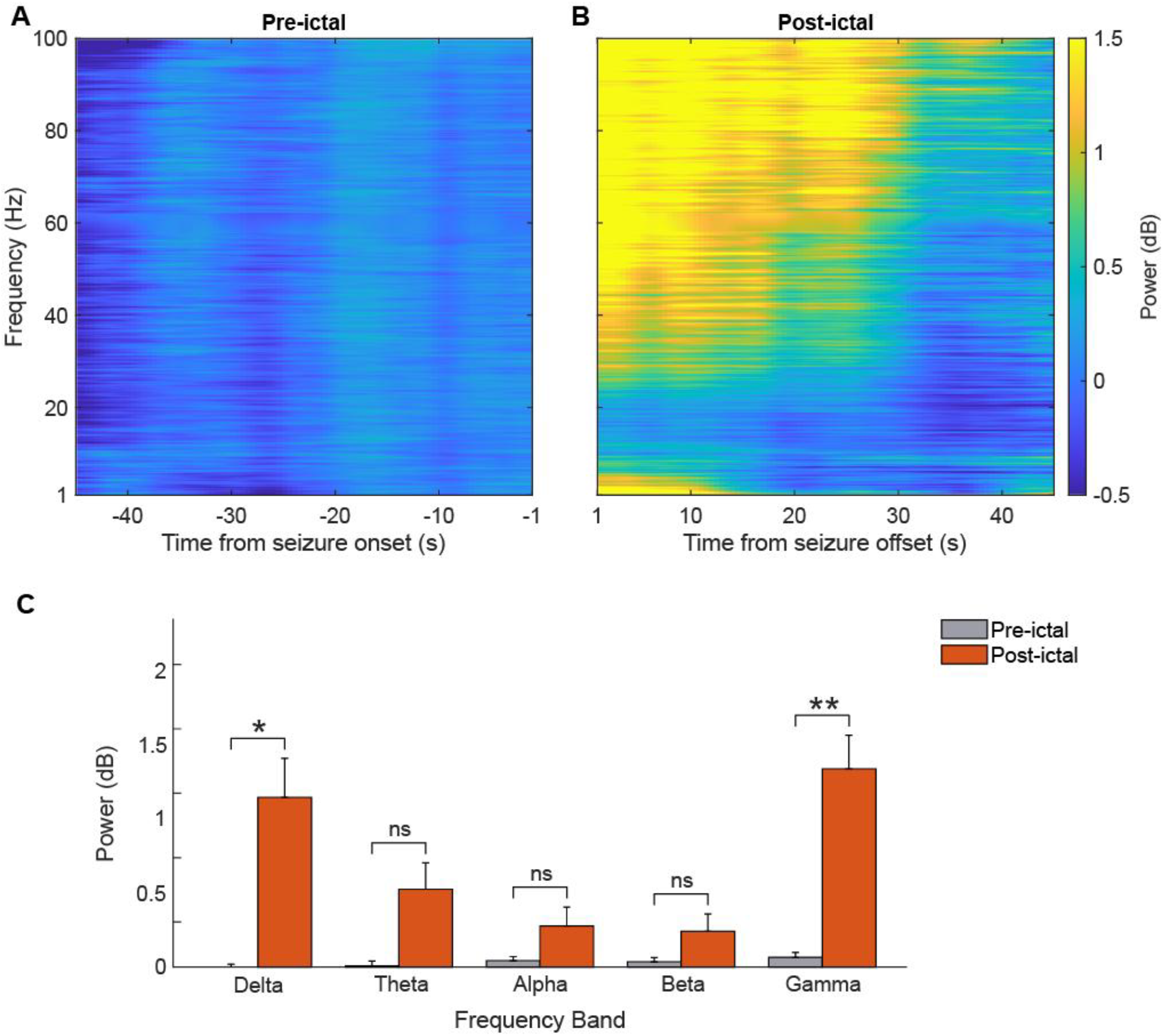
Transient increase in postictal EEG gamma and delta power in absence epilepsy. (**A, B**) Group-mean scalp-EEG power spectrograms expressed relative to each seizure’s own preictal baseline, shown separately for the preictal period **(A**, aligned to seizure onset) and the postictal period **(B**, aligned to seizure offset). Power was averaged across all EEG channels and across all 232 seizures in 55 patients and smoothed with a 10 s moving average. Preictal power remained relatively unchanged over time, whereas postictal power increased within the first seconds after offset, most prominently at higher and lower frequencies, and returned to baseline after 15 to 30 s. (**C**) EEG power averaged over the whole postictal window (1 to 45 s) versus the time-matched preictal window (-45 to -1 s) for delta (1 to 3 Hz), theta (4 to 7 Hz), alpha (8 to 13 Hz), beta (14 to 24 Hz) and gamma (25 to 100 Hz) frequency bands. Bars show mean ± SEM across seizures. Power was significantly increased postictally in the delta (+1.15 dB) and gamma (+1.34 dB) bands, but not in the theta, alpha, or beta bands. Data are from same patients as seizures as in **A, B**. Brackets denote paired Wilcoxon signed-rank tests with Benjamini-Hochberg false-discovery-rate correction across the five bands (*q < 0.05, **q < 0.01; ns, not significant), applied with an effect-size threshold of |d| ≥ 0.10.

## Discussion

We found that across 55 patients in four cohorts, absence seizures were followed by a brief, broadband increase in scalp EEG power, particularly in the delta and gamma bands. This characteristic physiological signature is consistent with cortical activation,^6-8^ and appeared within 1 s after offset, resolving by 15 to 30 seconds. These results imply that rather than being a passive return to baseline, emergence from an absence seizure is an active, structured event.

The presence of an arousal-like EEG signature at seizure offset supports the hypothesis that arousal systems contribute to the termination of absence seizures and facilitate recovery. If emergence is an active, arousal-like event, the ascending arousal circuitry that regulates thalamocortical excitability, including brainstem cholinergic and noradrenergic nuclei, the intralaminar thalamus, and widespread cortices, represents a plausible substrate. Arousal counteracts absence seizures across multiple time scales: increased arousal reduces absence seizure probability,^1; 2^ and salient sensory stimuli can interrupt ongoing seizures.^4; 5^ The observation of an arousal-like signature at spontaneous offset suggests that these systems may participate in the natural termination of absence seizures and the subsequent recovery of awareness, complementing network-based accounts of impaired consciousness in absence epilepsy.^9^

Previous work mainly investigated EEG or fMRI changes preceding absence seizures rather than the postictal period.^2; 9-12^ Of note, previous studies in rodent models reported EEG changes consistent with progressively decreasing arousal in the seconds prior to SWDs, ^10; 11^ which we did not observe in the current human data. This could be attributed to different arousal time courses in humans and rodent models, where the human arousal state may be reduced for much longer time periods prior to absence seizures,^1; 2^ well beyond the 45 s preictal window analyzed here. This possibility should be investigated further in future work using methods better suited to detecting subtle long-term changes in arousal state, such as measurements of pupil diameter or autonomic function. In addition, the rodent preictal signature was recorded by invasive electrocorticography, whereas in human scalp EEG the preictal signals may be attenuated or otherwise differ from rodents. Thus, the lack of preictal EEG changes in the present work does not preclude a human preictal change.

The present work suggests several other important directions for future investigations. Without simultaneous behavioral testing, the EEG signature observed in the postictal period cannot yet be linked to recovery of awareness. Thus, whether arousal drives seizure termination or merely follows it remains unresolved. Therefore, future studies should integrate postictal EEG with postictal reaction time or awareness assessments to determine whether the arousal signature corresponds to behavioral recovery. Source localization or simultaneous EEG-fMRI may help identify the underlying neural networks and structures involved in this process in a more meaningful way. Further investigations should also address whether augmenting postictal arousal, either pharmacologically, through neurostimulation or other means, accelerates seizure termination and recovery.

Taken together, these results suggest that arousal plays a role in recovery from absence seizures, which is an active electrophysiological event and not a passive return to rest. With further investigation, postictal arousal may serve as a marker and as a therapeutic target for recovery of consciousness in absence epilepsy.

## Data Availability

All data produced in the present study are available upon reasonable request to the authors.

## Ethical Publication Statement

We confirm that we have followed the Journal’s ethical guidelines for publication and that this report meets those standards.

